# Electrocardiographic Abnormalities and Troponin Elevation in COVID-19

**DOI:** 10.1101/2020.11.12.20230565

**Authors:** Ehud Chorin, Matthew Dai, Edward Kogan, Lalit Wadhwani, Eric Shulman, Charles Nadeau-Routhier, Robert Knotts, Roi Bar-Cohen, Chirag Barbhaiya, Anthony Aizer, Douglas Holmes, Scott Bernstein, Michael Spinelli, David Park, Larry Chinitz, Lior Jankelson

## Abstract

**Background:** the COVID19 pandemic has resulted in worldwide morbidity at unprecedented scale. Troponin elevation is a frequent laboratory finding in hospitalized patients with the disease, and may reflect direct vascular injury or nonspecific supply-demand imbalance. In this work, we assessed the correlation between different ranges of Troponin elevation, Electrocardiographic (ECG) abnormalities and mortality.

**Methods:** We retrospectively studied 204 consecutive patients hospitalized at NYU Langone Health with COVID19. Serial ECG tracings were evaluated in conjunction with laboratory data including Troponin. Mortality was analyzed in respect to the degree of Troponin elevation and the presence of ECG changes including ST elevation, ST depression or T wave inversion.

**Results:** Mortality increased in parallel with increase in Troponin elevation groups and reached 60% when Troponin was >1 ng/ml. In patients with mild Troponin rise (0.05 – 1.00 ng/ml) the presence of ECG abnormality resulted in significantly greater mortality.

**Conclusion:** ECG repolarization abnormalities may represent a marker of clinical severity in patients with mild elevation in Troponin values. This finding can be used to enhance risk stratification in patients hospitalized with COVID19.

## Introduction

Coronavirus Disease (COVID-19), caused by severe acute respiratory syndrome coronavirus-2 (SARS-CoV-2), is now one of the deadliest pandemics in modern history. [1]. As of September 15 2020, over 30 million individuals were reported to be infected by SARS-CoV-2, with more than 950,000 deaths [2]. Recent reports [3, 4] revealed that cardiac complications are common (≈20-25%) in COVID-19 infection and are associated with increased mortality. However, in those reports, “cardiac complications” were defined according to clinical and laboratory parameters (troponin levels) without systematic electrocardiographic (ECG) evaluation. Whether these troponin elevations represent primary myocardial infarction, supply-demand inequity, or non-ischemic direct myocardial injury remains unclear. ECG is a highly available diagnostic test that can be quickly performed without exposing a large number of personnel to SARS-CoV-2. ECG has demonstrated prognostic value in population-based studies [5, 6] and thus offers a particularly appealing modality during the current pandemic. We thus sought to determine whether findings on the first presenting ECG provide prognostic information and provide insights on myocardial injury. We reviewed ECGs of consecutive patients with COVID-19 infection requiring hospitalization. We examined our findings stratified by troponin levels and clinical condition.

## Methods

This is a retrospective study performed at NYU Langone Medical Center, New York, USA. We included 204 consecutive adult patients hospitalized at NYU Langone Medical Center with COVID-19 disease. Medical records were reviewed to obtain baseline characteristics, laboratory data, and serial ECGs. Troponin I concentrations were assessed via the Abbott Architect method (Abbott, Abbott Park, Illinois) wherein the 99th percentile for a normal population is 0.05 ng/mL. Descriptive analyses were performed by troponin levels stratified into normal (0.00-0.05 ng/mL), mildly elevated (0.05-1 ng/mL), and significantly elevated (>1 ng/mL). ECGs were reviewed and interpreted by five seniors cardiologists who were blinded to the clinical status of the patients. Data reviewed from each ECG included heart rate, rhythm categorized as normal sinus rhythm or atrial fibrillation/flutter (AF), atrioventricular block (AVB), right bundle branch block (RBBB), left bundle branch block (LBBB), a nonspecific intraventricular conduction block (QRS duration >120ms), the presence of ST segment or T-wave changes (localized ST elevation, localized T-wave inversion, or other nonspecific repolarization abnormalities). The closing date of follow-up was April 15th 2020. Collected data on the closing date included arrhythmic events and mortality. The study was reviewed and approved by the NYU Institutional Review Board and Quality Improvement initiative in accordance with the ethical standards laid down in the 1964 Declaration of Helsinki and its later amendments, with a waiver of informed consent.

### Statistical analysis

Statistical analysis was performed using IBM SPSS Statistics 26, and figures were constructed using GraphPad Prism 8. Continuous variables are expressed as mean ± standard deviation, and categorical variables are expressed as count (percentages). Normality of data samples was assessed using Shapiro-Wilk test. Two sample hypothesis testing for continuous variables was performed using Student’s *t*-test if samples had normal distributions and Mann-Whitney *U* test if samples did not have normal distributions. Hypothesis testing for categorical variables was performed using Fisher’s exact test. Significance testing for Kaplan-Meier curves was performed using log-rank test. For predictors of mortality, univariate analysis was performed using Cox proportional hazards regression, and significant univariate predictors were included in the multivariate analysis.

## Results

We included 204 patients in our cohort with a mean follow up time of 24.2 ± 7.4 days. The clinical and epidemiological characteristics stratified by ECG abnormalities are presented in Table 1. The mean age was 64±13 years and 76% were male. Comorbidities were common: 30% of patients had diabetes mellitus, 56% had hypertension, 12% had coronary artery disease, 3% had heart failure, and 6% had COPD. Baseline electrocardiographic characteristics revealed mean HR of 89 ± 16 bpm and mean Bazett-corrected QT interval of 444 ± 26 ms. The vast majority were in normal sinus rhythm (95%), while 5% of patients had AF. Atrioventricular block was rare: 9 (4%) patients had a first degree block and no patients had second or third degree AV block. Abnormal intraventricular conduction was found in 11% (with RBBB in 8%, LBBB in 3%). Repolarization abnormalities (ST elevation, ST depression or T wave inversion) were common (36 patients, 17.6%): one patient (0.5%) had localized ST elevation, 12 (5.9%) had ST depression and 28 (13.7%) had localized T-wave inversion. Patients with repolarization abnormalities demonstrated higher troponin levels and a trend towards higher mortality (Table 1). One patient presented with a fever of 103.1F which unmasked a previously unknown type I Brugada pattern (Figure 1). Fifty (25%) patients died of respiratory or multi-organ failure. In univariate and multivariate Cox regression analyses, clinical predictors of death included age and elevated Troponin (Table 2), but did not include gender, race or cardiovascular comorbidities (CAD, CHF, HTN). The mortality rate increased with incrementally higher troponin group: 14/120 [11.7%] for patients with negative troponin, 24/64 [37.5%] for patients with mildly elevated troponin and 12/20 [60%], for patients with significantly elevated troponin (p < 0.01), Figure 2. The presence of an abnormal ECG finding resulted in significantly lower survival in the intermediate Troponin elevation group (0.05-1 ng/ml) but not in the low (<0.05 ng/ml) or high (> 1 ng/ml) Troponin elevation groups (Figure 3).

**Table 1.**
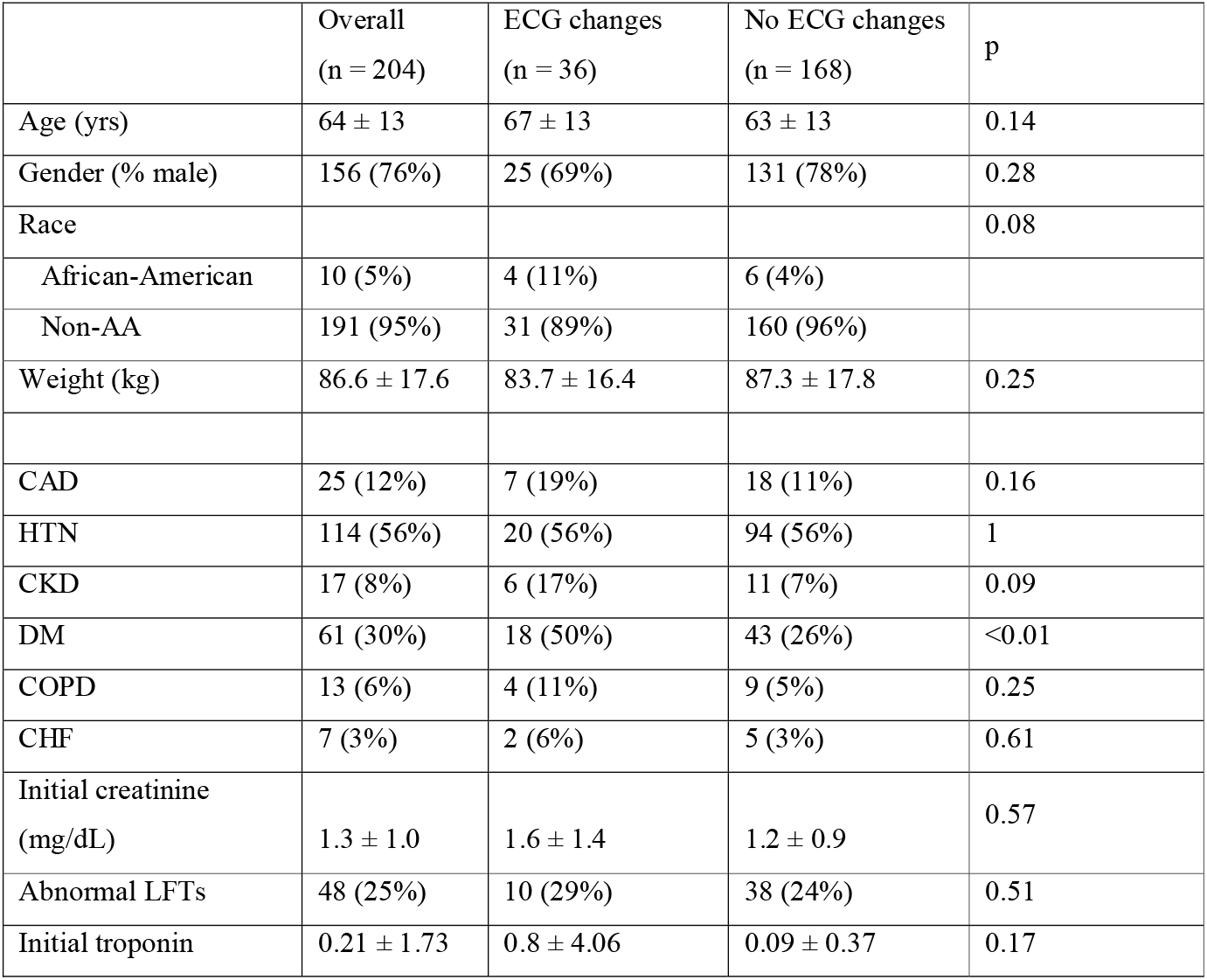
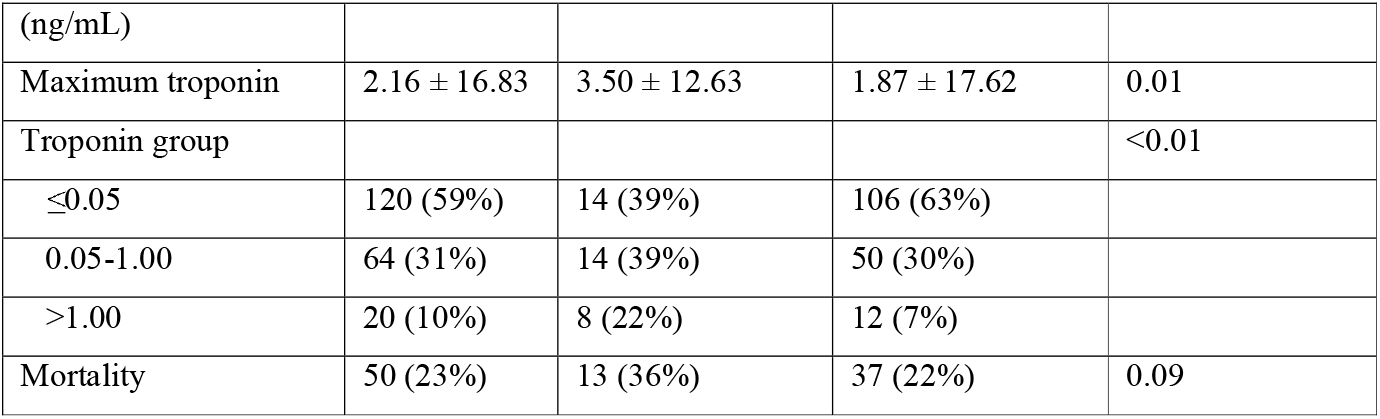
Baseline Characteristics

**Table 2.** Predictors of Mortality

| Univariate Regressions |  |  |  |
| --- | --- | --- | --- |
| Variable | HR | 95% CI | *p* value |
| Age (yrs) | 1.08 | 1.05 - 1.1 | <0.01 |
| Gender (% male) | 1.09 | .56 - 2.13 | 0.80 |
| Race |  |  |  |
| African-American | 0.8 | .2 - 3.3 | 0.76 |
| Weight (kg) | 0.99 | .97 - 1.01 | 0.18 |
| CAD | 1.08 | .49 - 2.41 | 0.84 |
| HTN | 1.39 | .78 - 2.48 | 0.26 |
| CKD | 2.4 | 1.12 - 5.11 | 0.02 |
| DM | 1.77 | 1.01 - 3.1 | 0.05 |
| COPD | 1.46 | .53 - 4.07 | 0.47 |
| CHF | 2.09 | .65 - 6.73 | 0.22 |
| Initial creatinine (mg/dL) | 1.24 | 1.08 - 1.43 | <0.01 |
| Abnormal LFTs | 0.84 | .42 - 1.7 | 0.64 |
| Initial troponin (ng/mL) | 0.99 | .84 - 1.16 | 0.91 |
| Maximum troponin | 1.01 | 1. - 1.02 | 0.02 |
| Positive troponin (>0.05) | 4.24 | 2.28 - 7.86 | <0.01 |

| Multivariate Regression |  |  |  |
| --- | --- | --- | --- |
| Variable | HR | 95% CI | *p* value |
| Age (yrs) | 1.06 | 1.04 - 1.1 | <0.01 |
| CKD | 1.53 | .65 - 3.6 | 0.33 |
| DM | 0.99 | .53 - 1.86 | 0.98 |
| Positive troponin (>0.05) | 3.22 | 1.71 - 6.05 | <0.01 |

**Figure 1:**
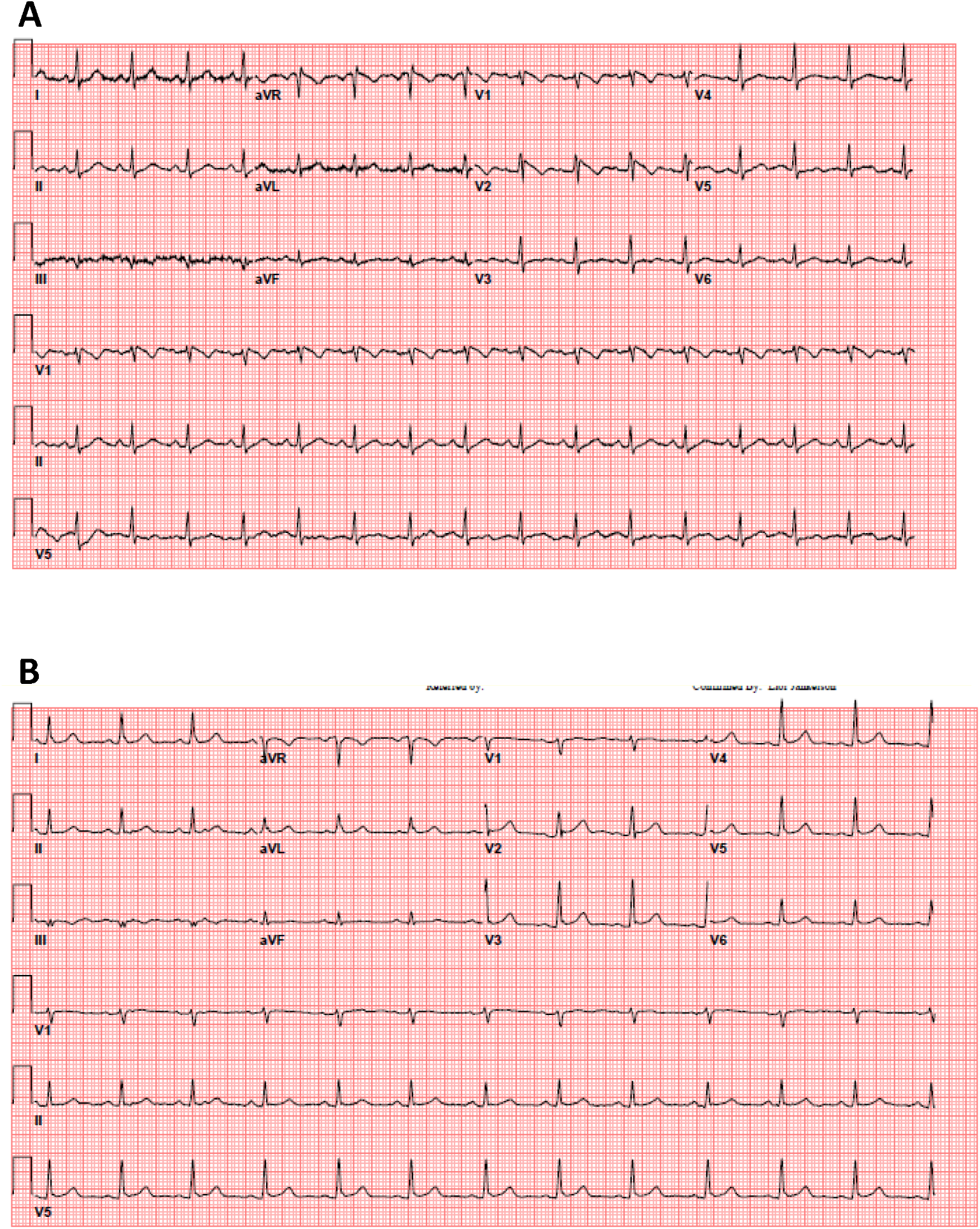
35 year old female patient without significant medical history presented with a fever of 103.1 F. A. The patient’s initial 12-lead electrocardiogram in the emergency department. B. The patient’s repeat 12-lead electrocardiogram with resolution of fever.

**Figure 2:**
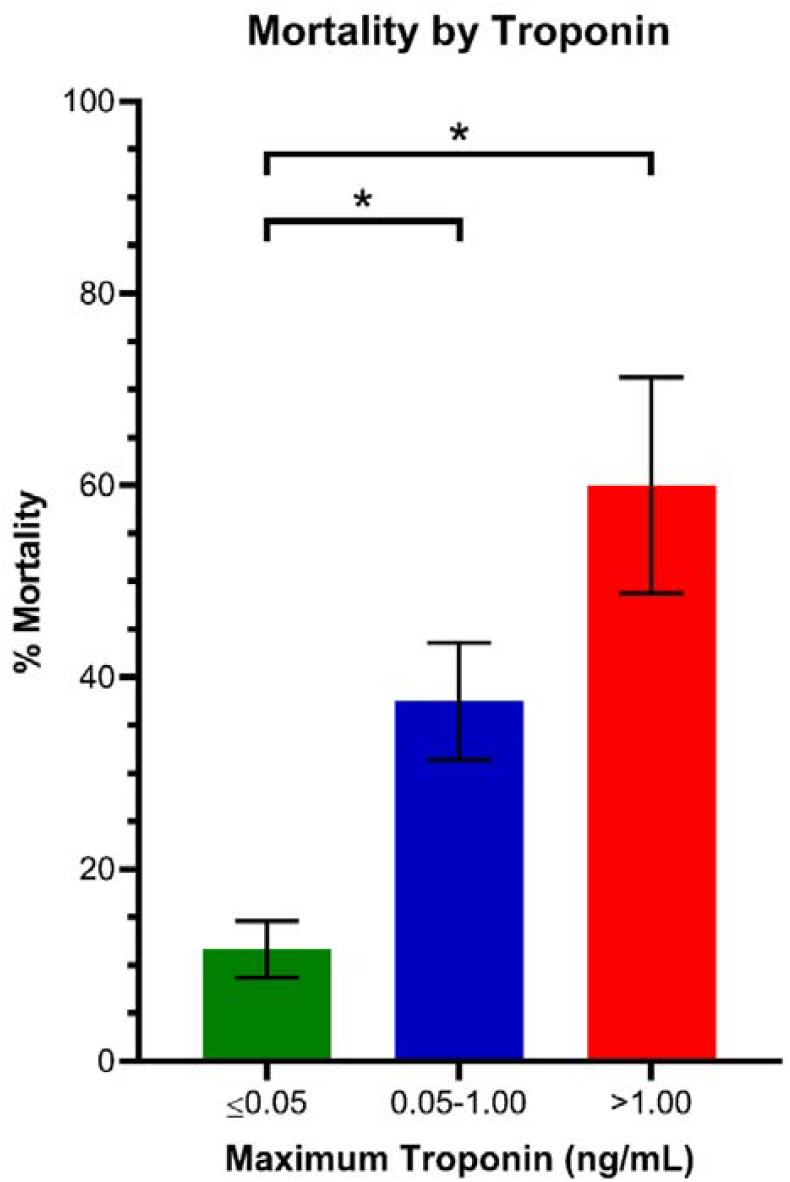
Mortality rate stratified by Troponin elevation groups. * P<0.01

**Figure 3:**
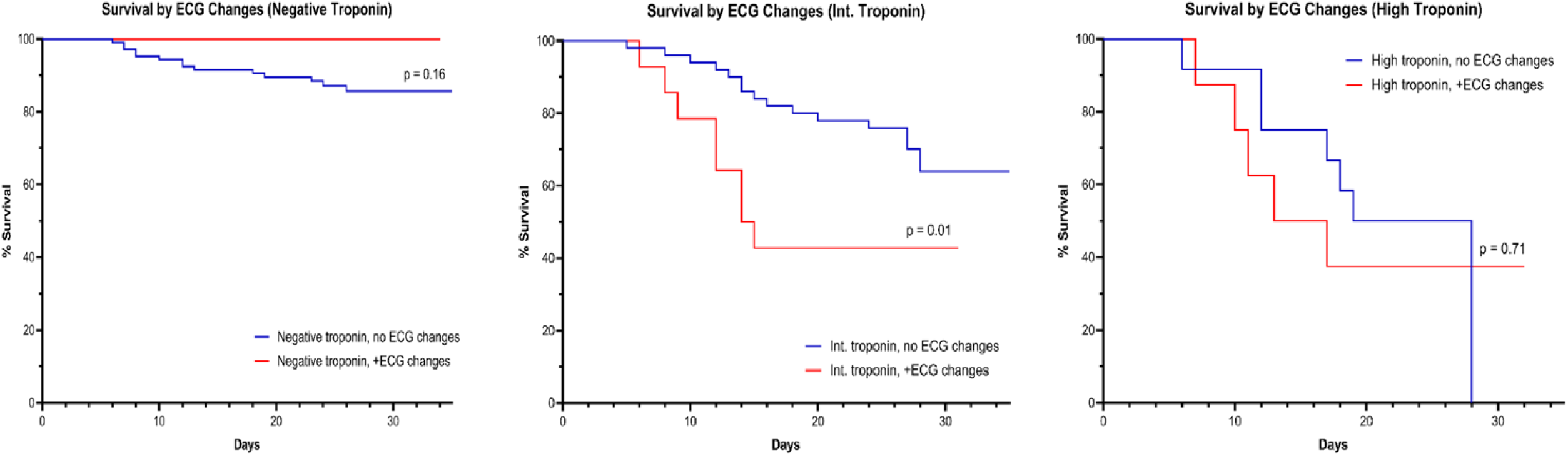
KM survival according to ECG changes stratified by Troponin elevation group

## Discussion

COVID-19 has been shown to cause cardiovascular morbidity by direct myocardial injury as a result of the inflammatory cascade or cytokine release, microvascular damage due to disseminated intravascular coagulation and thrombosis, direct entry of SARS-CoV-2 into myocardial cells via binding to ACE2 receptors, and hypoxemia combined with increased metabolic demands of acute illness leading to myocardial injury [9-11]. In this retrospective cohort study we further assess the interaction of ECG abnormalities and Troponin elevation. We demonstrate that 1) myocardial injury defined by elevated Troponin is common among patients hospitalized with COVID-19 but is more often mild, associated with low-level elevation in troponin concentration. 2) more significant myocardial injury, as evident by increased Troponin level may be associated with higher risk of mortality. 3) In the group of patients with mild Troponin elevation (0.05-1 ng/ml), ECG abnormalities are associated with significantly increased mortality.

Though troponin elevation above the 99th percentile of the upper reference limit is considered the central marker of “myocardial injury”, mild elevation between 0.05 to 1 is often nonspecific and associated with non-vascular etiologies such as strain, myocyte necrosis and increased cell membrane permeability [12]. Indeed, mild Troponin elevation was a frequent finding in our cohort, present in 31% of patients with COVID19. In this regard, our data suggests that assessment for the presence of ECG abnormalities can be used to enhance inpatient risk stratification in those patients with mild Troponin elevation. Finally, as persistent fever is a frequent clinical feature of COVID-19, caregivers should be familiar with the phenomena of fever induced Brugada pattern and not mistake it for ST elevation myocardial infarction.

### Limitations

Our study has several limitations. This is an observational, retrospective study. Because of its retrospective nature, the study is subject to selection bias, and its results imply association, not cause and effect. Relatively short follow-up time after was available.

## Data Availability

review of request will be performed

